# Comparing ancestry calibration approaches for a trans-ancestry colorectal cancer polygenic risk score

**DOI:** 10.1101/2023.10.23.23296753

**Authors:** Elisabeth A. Rosenthal, Li Hsu, Minta Thomas, Ulrike Peters, Christopher Kachulis, Karynne Patterson, Gail P. Jarvik

## Abstract

**Background:** Colorectal cancer (CRC) is a complex disease with monogenic, polygenic and environmental risk factors. Polygenic risk scores (PRS) are being developed to identify high polygenic risk individuals. Due to differences in genetic background, PRS distributions vary by ancestry, necessitating calibration.

**Methods:** We compared four calibration methods using the All of Us Research Program Whole Genome Sequence data for a CRC PRS previously developed in participants of European and East Asian ancestry. The methods contrasted results from linear models with A) the entire data set or an ancestrally diverse training set AND B) covariates including principal components of ancestry or admixture. Calibration with the training set adjusted the variance in addition to the mean.

**Results:** All methods performed similarly within ancestry with OR (95% C.I.) per s.d. change in PRS: African 1.5 (1.02, 2.08), Admixed American 2.2 (1.27, 3.85), European 1.6 (1.43, 1.89), and Middle Eastern 1.1 (0.71, 1.63). Using admixture and an ancestrally diverse training set provided distributions closest to standard Normal with accurate upper tail frequencies.

**Conclusion:** Although the PRS is predictive of CRC risk for most ancestries, its performance varies by ancestry. *Post-hoc* calibration preserves the risk prediction within ancestries. Training a calibration model on ancestrally diverse participants to adjust both the mean and variance of the PRS, using admixture as covariates, created standard Normal z-scores. These z-scores can be used to identify patients at high polygenic risk, and can be incorporated into comprehensive risk scores including other known risk factors, allowing for more precise risk estimates.

## Introduction

Colorectal cancer (CRC; MIM: 114500) is the third most common cancer in the United States (U.S.), and is associated with the second most deaths from cancer ^1^. CRC risk may be attributable to social determinants of health such as socioeconomic status, health care access, and food security, which are known to be associated with race ^2^. Individuals who self-report as Black or Native American/Alaskan Native have the highest lifetime risk for CRC as well as higher mortality rates ^1,3,4^. Additionally, risk for CRC is affected by other environmental factors such as diet, smoking and physical activity, as well as genetics ^5^. Although 25% of CRC appear to be familial, only 20% of familial CRC is explained by variation at single genes ^6–8^.

In aggregate, low risk SNPs across the genome also contribute to risk of CRC. Polygenic risk scores (PRS), which aggregate the effect of genetic variants across the genome, are currently being developed to aid in identifying individuals at higher genetic risk of developing CRC ^9,10^. As these PRS have been shown to be independent of family history, they may provide orthogonal information when incorporated into a comprehensive score ^11^. Additionally, PRS have been shown to be more predictive at younger ages, making them useful for identifying patients who can benefit from increased or earlier screening, as current environmental risk prediction models are optimized for middle-aged patients ^12,13^.

Development of PRS involves estimation of SNP effects, which are influenced by the genetic ancestry of the cohorts used to develop the PRS through linkage disequilibrium (LD) across the genome and minor allele frequency (MAF) at the SNPs^14^. The PRS for CRC used here (PGS catalog PGS003852)^15^, was developed with participants of European and East Asian ancestry, using Bayesian methods that estimate SNP effect sizes accounting for LD in both populations, while simultaneously estimating the number of causal SNPs given the heritability of CRC ^9,16^. When participants from multiple ancestries are included, this method helps delineate the collinearity of SNPs in LD and therefore improves identification of SNPs that are associated with CRC risk in multiple ancestries, reporting a single estimated effect for each SNP ^17,18^.

Even when more than one genetic ancestry is used to develop a multi-ancestry PRS, the distribution of the PRS will differ by genetic ancestry, as LD and MAF vary across genetic ancestries. Therefore, an individual’s PRS must be interpreted within the context of their unique genetic ancestry in order to estimate their PRS-specific predicted genetic risk. However, genetic ancestry is not strictly categorical, due to admixture, and cannot be adequately determined by observable phenotypes in the clinic, such as skin color, hair type, or eye shape ^19,20^. Several *post-hoc* mechanisms for calibrating the PRS for genetic ancestry in the clinic setting have been proposed ^21,22^. These include using linear models, based on a training data set of diverse populations, to adjust the mean PRS using principal components (PCs) of ancestry. Additionally, methods using training models have been adapted to adjust for both the expected mean and variance of the PRS. One benefit of these approaches is that patients do not need to be categorized within a genetic ancestry. Furthermore, these approaches can be applied to admixed individuals, who make up an increasing proportion of U.S. residents.

We set out to determine which, if any, of these *post-hoc* PRS calibration methods is preferable in the context of this multi-ancestry CRC PRS, by analyzing data from the All of Us Research Program (AOU)^23,24^. The purpose of AOU is to collect survey, electronic health record (EHR), and genotype data on diverse participants who live in the U.S., for use in broad research and to help reduce inequities in healthcare in the U.S.. As this data set contains both ancestrally diverse and admixed participants, it is an ideal biobank to assess *post-hoc* genetic ancestry calibration methods for use in the clinic.

## Subjects and Methods

### Case/Control assignment

We updated a previously developed CRC case/control algorithm (see Web Resources) to define CRC cases and controls. This algorithm was created using International Classification of Disease (ICD) 9 codes and Current Procedural Terminology (CPT) codes. The AOU dataset contains a rich database of ICD 9 and 10 codes, CPT codes, and Logical Observation Identifiers Names and Codes (LOINC). As the completeness of this data varies among participants^25^ we attempted to widen our net by creating concept sets based on the original ICD 9 and CPT codes, incorporating the related ICD 10 codes and LOINC codes. Each concept set relates to a different table from the word files provided by the algorithm’s authors (Supplemental Tables S2-S10). The algorithm was developed to screen for CRC cases with potentially monogenic causes of CRC, and therefore excluded individuals with ulcerative colitis (UC) or Crohn’s disease, two disorders associated with CRC. As we are interested in a polygenic component underlying CRC we did not exclude participants with UC or Crohn’s disease. Additionally, we excluded all participants with a known monogenic (pathogenic or likely pathogenic variant in ClinVar) cause of CRC in any of *AKT1* [MIM: 164730], *APC* [MIM: 611731], *AXIN2* [MIM: 604025], *BMPR1A* [MIM: 601299], *CDH1* [MIM: 192090], *CHEK2* [MIM: 604373], *CTNNA1* [MIM: 116805], *EPCAM* [MIM: 185535], *GALNT12* [MIM: 610290], *GREM1* [MIM: 603054], *MLH1* [MIM: 120436], *MSH2* [MIM: 609309], *MSH3* [MIM: 600887], *MSH6* [MIM: 600678], *MUTYH* [MIM: 604933], *NTHL1* [MIM: 602656], *PDGFRA* [MIM: 173490], *PIK3CA* [MIM: 171834], *PMS2* [MIM: 600259], *POLD1* [MIM: 174761], *POLE* [MIM: 174762], *PTEN* [MIM: 601728], *RPS20* [MIM: 603682], *SMAD4* [MIM: 600993], *STK11* [MIM: 602216], or *TP53* [MIM: 191170] ^24^ *(in press)*.

Potential cases included all participants that had at least one CRC diagnosis in their medical record (Supplemental table S2) but did not have a known pathogenic variant for CRC or a diagnosis for monogenic disorders with increased risk of CRC (Supplemental table S3). The cleaned set of cases was derived from potential cases using the following ordered algorithm:

A. They had a surgical procedure related to CRC within a year of diagnosis (Supplemental Table S4)
B. If not (A), then they had chemotherapy or radiation (Supplemental Tables S5 and S6) within a year of CRC diagnosis <u>and</u> they did not have other types of cancers listed in the exclusion table (Supplemental Table S7)
C. If not (A) or (B) and they had at least two CRC diagnosis codes within 2 years of each other and no other cancers listed in the exclusion table

Potential controls were participants with no diagnosis codes for CRC or a monogenic disorder with increased risk of CRC, and no evidence of a pathogenic variant for CRC. Using guidance from the algorithm, the screened controls met either of the following criteria:

A. They had at least one sigmoidoscopy or colonoscopy and no positive pathology report (Supplemental Table S8)
B. They had at least two instances of a fecal immunochemical test (FIT) or a fecal occult blood test (FOBT) that were at least 5 years apart and never had a positive lab result from any of these tests (Supplemental Tables S9 and S10)

All other participants that were not assigned case/control status, and were not excluded by the algorithm, were left unassigned. Unassigned participants are included in the analysis except when statistical testing involves case and control status. Some analyses include age and sex. Age is defined as observational age: age at onset of cases, age at last screening for controls, and age at consent to AOU for the unassigned participants. In these analyses we further restricted analysis to participants who were older than the minimum observed age of onset of CRC, 19, as CRC rarely presents so early. Sex is defined as sex assigned at birth.

### Genetic Ancestry Clustering

We calculated 32 genome-wide PCs of ancestry using an unrelated subset of reference global genomes from 1000 Genomes (1KG) and Human Genome Diversity Project (HGDP) (N = 4,151)^26,27^. We then projected the AOU participants onto the PC space. We used EIGMIX to estimate admixture in the AOU participants for the continental ancestry groups represented by the reference genomes: African (AFR), Admixed American/Latino (AMR), East Asian (EAS), European (EUR), Middle Eastern (MID) and South Asian (SAS)^28^. See supplemental methods for details on PC and admixture calculation. We then clustered the AOU participants into ancestry groups, using the estimated percent ancestries, in order to compare the performance of the PRS across continental ancestry groups. As the amount of genetic diversity among different racial and ethnic groups in the United States differs (Supplemental Figures S1 and S2)^28–30^, we used an 80% cutoff to cluster individuals into EAS, EUR and SAS clusters, and a 60% cutoff to cluster individuals into AFR, AMR and MID clusters. All other participants are referred to as Other (OTH). A subset of highly admixed participants from OTH, called ADM, contains participants with <50% estimated ancestry for all continental ancestries considered.

### PRS calculation

Details of PRS calculation in AOU and the reference 1KG and HGDP data are given in the supplemental methods. In brief, we extracted genotype data from WGS using HAIL^31^ on the Spark cluster. After performing quality control, we calculated the PRS as the sum of the genotype effects, *PRS_j =_* 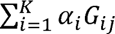 for each participant j, where *G*_ij_ is the count of the affect allele at SNP i for participant j, α_i_ is the allele affect for SNP i, and K is the total number of SNPs, using the R package bigSNPR v. 1.10.8. The allele effects for each SNP were obtained from the corresponding author of PMID: 36789420 ^9^ and can also be found in the PGS catalog (PGS003852).

### PRS Calibration

As the raw PRS follows a Normal distribution within genetic ancestries, it is reasonable to calibrate to the standard Normal distribution. Therefore, all calibration methods considered here follow the general standardization protocol to calculate the adjusted score PRS_Adj,j_ for individual j:

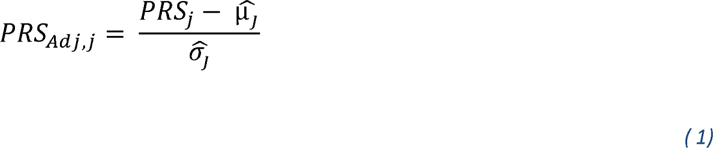

where 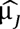 is the expected mean and 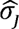 is the expected standard deviation (s.d.) for individual j. The calculated expected mean and s.d. depend on the calibration method.

We started with three *post-hoc* calibration methods of the raw PRS (RAW) to account for ancestry: linear model to adjust for the first 5 PCs of ancestry in AOU (PC_μ), linear model to adjust for the estimated admixture percentages (AD) in AOU (AD_μ), and a previously published method which adjusts both mean and s.d. as a function of PCs using a trained model on a reference dataset (PC.REF_μσ)^21,22^. The linear model calibrations (PC_μ and AD_μ) are based on regressing the PRS on the covariates, 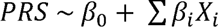, in the AOU dataset, so that the expected mean for individual j is calculated as 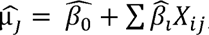, where 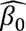 is the intercept term from the linear model and 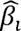 is the estimated effect of covariate X_i_, where X are the first five PCs of ancestry (PC_μ) or the admixture estimates (AD_μ). The expected s.d. for each individual is calculated over all samples in AOU, 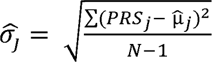, where N is the sample size. The ancestry adjusted PRS based on PC_μ and AD_μ calibration methods are given below:

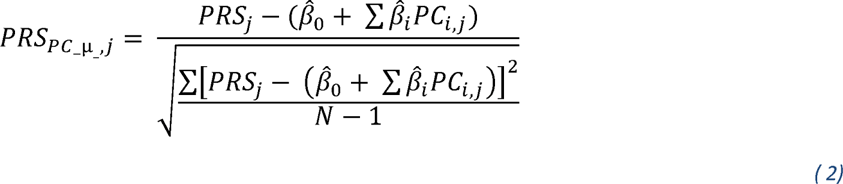

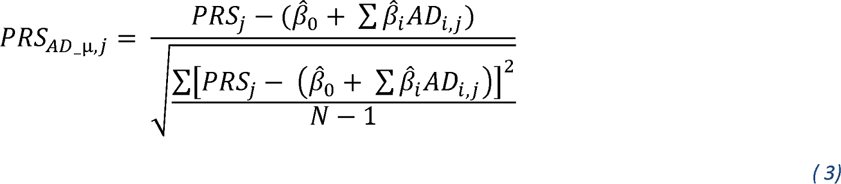

Similarly, calibration using a training data set, calculates the expected mean for each individual in AOU using the regression coefficients 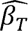 calculated only on the training data set, T. In addition, the s.d. is estimated by regressing the residual variance, δ, on the covariates in the training set 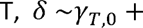 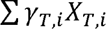, so that 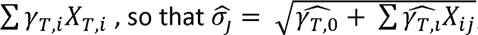, for each individual j, where the covariates, X_T_, are the first 5 PCs of ancestry in the training data and X_j_, are the first 5 PCs of ancestry for individual j in AOU. In this case, the training data set is the 1KG and HGDP reference genomes.

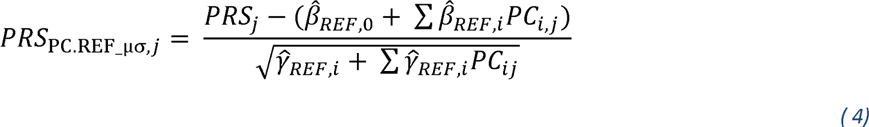

We chose to use 5 PCs of ancestry to make comparisons fair with AD_μ which has 5 degrees of freedom due to the six global ancestries considered in this analysis. We added in two more adjustments after observing the distribution of PC.REF_μσ. We modified the method by using a different training set randomly sampled from the unassigned participants in AOU, called AOU.REF, as they more closely resemble the cases and controls in AOU. The ancestry adjustment based on the modified calibration method, PC.AOU_μσ, is calculated as below:

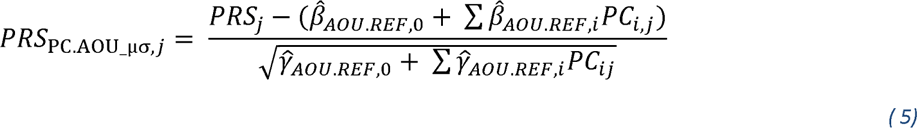

We also created a calibration model AD.AOU_μσ using the admixture estimates in AOU.REF, as given below.

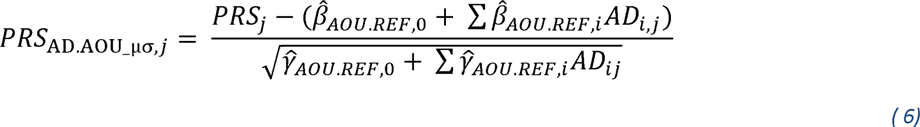

The properties of the calibration methods are given in Table 3.

We kept the patient reference sample size for AOU.REF (N=4,151) the same as the global reference sample size (1KG and HGDP) so that we could make fair comparisons. As the AOU dataset is heavily weighted toward individuals of EUR ancestry, a simple random sample would also be heavily weighted in this way. Therefore, we attempted to randomly sample from the space of the first 5 PCs. First, we calculated the geometric median of the first five PCs. Second, we calculated the Euclidean distance to this median for each participant. We divided the distances into five equal ranges. We then randomly sampled an equal number of participants from each range, resulting in the following counts of unassigned participants from each ancestry cluster in the reference patient sample: 1108 AFR; 1459 AMR; 84 EAS; 820 EUR; 72 MID; 577 OTH; 31 SAS.

### Comparing raw and calibrated PRSs

We evaluate the calibration methods by their ability to minimize variation in the PRS that is due to genetic ancestry unrelated to CRC. That is, the calibration should result in the same distribution regardless of genetic ancestry so that all values are on the same scale. As mentioned above, the calibrations are expected to result in a N(0,1) distribution. Therefore, we compare the overall mean and s.d. within genetic ancestries, across the calibration methods to their expectation of 0 and 1, respectively. Furthermore, we perform Kolmogorov-Smirnov (KS) tests for each calibration within ancestry. Rejecting the null hypothesis indicates that the distribution is not standard Normal. We used a conservative p-value cutoff of 0.05 for these tests, without adjusting for multiple testing. Failure to reject the null hypothesis suggests that the adjusted scores align with z-scores from a standard Normal distribution.

The calibration method should also preserve the effect size and significance that is observed for the association between the raw score with CRC. As these calibration methods are linear transformations of the raw score, they should not alter the observed association within ancestry clusters. Therefore, we compared three metrics. First, we compared the odds ratio (OR) for a single s.d. change in the score, with and without adjusting for age and sex, within each ancestry cluster. The OR measures if the score is associated with CRC risk. Second, we compared the log(OR) for each quintile compared to the middle quintile to determine if those at high risk (top quintile) can be differentiated from those at typical risk (middle quintile) within ancestry clusters, as suggested in. Third, we compared the area under the receiver operating curve (AUC), with and without adjusting for age and sex, within each ancestry cluster. The AUC compares how well the scores can distinguish between cases and controls, overall. We used 500 bootstrap iterations, with replacement, to estimate the 95% confidence intervals (C.I.) for the AUC.

Lastly, we compared the observed and expected upper 5, 7.5 and 10^th^ percentiles as these values could be used to identify patients at higher genetic risk for developing disease, relative to those of similar genetic ancestry. It is possible that the calibrations could result in distributions with skewed or heavy tails, which would result in too many or too few patients identified as being at higher risk. Observed percentiles were defined as the proportion of participants whose adjusted PRS was greater than the standard normal cutoffs (z=1.28, 1.44, or 1.64). The C.I. for the percentiles was calculated using the R function binom.test(). A C.I. that covers the expected percentile indicates that the upper tail is not skewed or heavy.

## Results

### Case/Control Demographics and Ancestry

We excluded 8 CRC affected participants who had penetrant Lynch syndrome or Familial Adenomatous Polyposis reported in their EHR and 455 participants with a known pathogenic or likely pathogenic variant for these monogenic disorders ^24^ *(in press)*. There were 668 participants remaining with at least one CRC code in the EHR, 348 of whom were classified as cases by the algorithm (Table 1, Supplemental Tables S2 and S3). There were 97,588 participants who had no evidence for CRC. Of these 12,378 had been screened for CRC with negative results and were assigned as controls. Using the ancestry clustering rule, the majority of participants clustered with EUR ancestry (45%), followed by AFR (23%), OTH (16%), AMR (9.5%), and MID (4.1)% with EAS and SAS making up < 3% of the sample (Table 1). As the sample size for EAS and SAS was small, we do not report statistical tests for these ancestries.

**Table 1:**
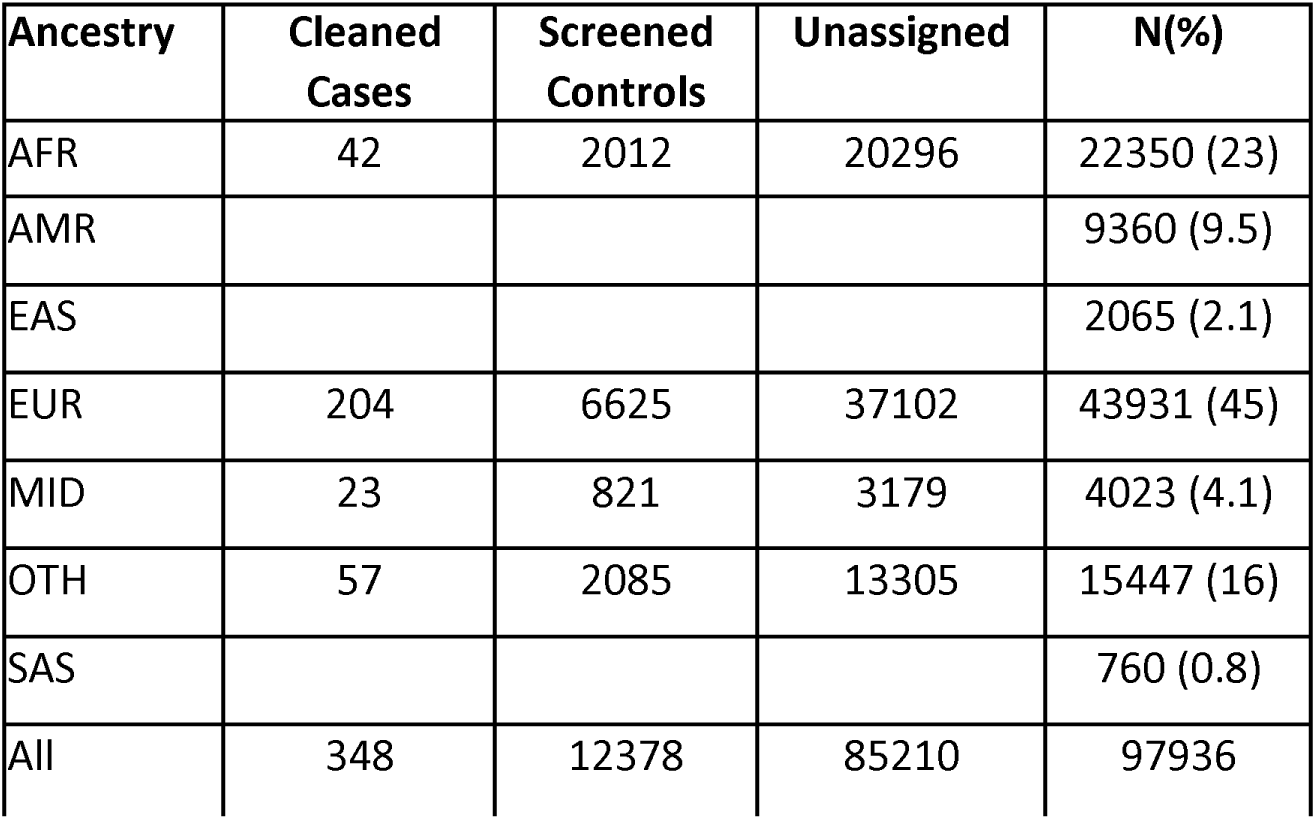
Count (percent) by ancestry and case status. “Cleaned cases” refers to participants assigned case status according to the CRC phenotyping algorithm. “Screened controls” refers to participants assigned control status by the CRC phenotyping algorithm. “Unassigned” refers to participants who were not excluded from analysis and were not assigned to case or control status by the CRC phenotype algorithm. Ancestries are AFR=African, AMR=Admixed American, EAS=East Asian, EUR=European, MID=Middle Eastern, OTH=Other, SAS=South Asian. For AMR, EAS and SAS, only total counts are given as the number of cases was <20, to comply with AOU publication rules.

The mean observed age of unassigned participants was about 10 years younger than that of the cases, overall (Table 2). The mean censored age of screened controls was similar to that of the cases, consistent with recommendations for screening in older patients. The observed age for both the cases and screened controls tended to be younger than their age at consent, indicating that most cases had CRC before entering the study and most screened controls were screened before entering the study.

**Table 2:** Mean observed age (min, max) by ancestry within all cleaned cases (age of onset), screened controls (age of last negative screening) and unassigned participants (age of consent). AFR=African, AMR=Admixed American, EAS=East Asian, EUR=European, MID=Middle Eastern, OTH=Other, SAS=South Asian. Age diff = difference between age of consent and observed age. Distributions are not reported for AMR, EAS and SAS due to AOU publication rules for sample sizes < 20.

| Ancestry | Cleaned cases | Screened controls | Unscreened controls |
| --- | --- | --- | --- |
| AFR | 57(41,83) | 57(18,90) | 48(18,104) |
| EUR | 62(26,89) | 61(18,94) | 54(18,103) |
| MID | 65(34,84) | 64(18,96) | 59(18,101) |
| OTH | 55(19,83) | 58(18,88) | 46(18,100) |
| All | 60(19,89) | 60(18,96) | 50(18,104) |
| Age diff | -5.2 (-27,2.5) | -2.5 (-34, 3.7) | 0 |

**Table 3:** Properties of the calibrations methods. The training data included either global reference samples (1KG and HGDP, N=4,151), all of All of Us participants (AOU, N=97,936) or a reference ancestrally diverse subset of AOU (AOU.REF, N=4,151). 1KG=1000 Genomes; HGDP=Human Genome Diversity Project; PC=Principal component of ancestry.

| Method | Training Data | Ancestry Variables | Variance Adjustment |
| --- | --- | --- | --- |
| **PC_μ** | AOU | PC1-PC5 | overall variance |
| **AD_μ** | AOU | Admixture estimates | overall variance |
| **PC.REF_μσ** | 1KG and HGDP | PC1-PC5 | linear model on PC1-PC5 |
| **PC.AOU_μσ** | AOU.REF | PC1-PC5 | linear model on PC1-PC5 |
| **AD.AOU_μσ** | AOU.REF | Admixture estimates | linear model on Admixture estimates |

Overall, males made up 49% of the cases and 40% of the controls. This sex difference may reflect sex differences in seeking healthcare and/or enrolling in genetic research studies. For AMR, EUR and OTH, the sex ratio difference was ≤ 10 percentage points. The sex ratio differences were > 20 percentage points for AFR and MID. We do not report exact counts in order to comply with AOU publication rules for cell counts less than 20.

### PRS Distribution Comparison

The distributions of RAW differed by ancestry in both their mean and s.d. (Table 4, Figure 1) due to differences in MAF and LD, as expected. The mean ranged between 0.05 (MID) and 0.4 (AFR) and the s.d. ranged between 0.41 (AFR) and 0.47 (most ancestries). The PC_μ means were close to zero for AFR, AMR and EAS, above zero for EUR and SAS, and below zero for MID and OTH. The s.d. for PC_μ was near 1 for all but AFR, which was below one. The KS test was not rejected for only EAS. AD_μ resulted in means closer to zero, but is above zero for MID. The s.d. for AD_μ is near 1 for all clusters, but is below 1 for AFR. The KS test was not rejected for AMR, EAS and SAS. Although the s.d. for PC.REF_μσ were nearly 1 for all clusters, PC.REF_μσ resulted in non-zero means for EAS, EUR, MID and SAS, prompting us to develop PC.AOU_μσ and AD.AOU_μσ. PC.AOU_μσ performed similarly to PC_μ. However the s.d. for AFR was closer to 1 and the s.d. for EAS, EUR and MID was slightly less than 1, and the KS test was not rejected for AFR. AD.AOU_μσ performed similarly to AD_μ. However, the s.d. for AD.AOU_μσ is < 1 for MID and EAS and > 1 for SAS. Furthermore, AD.AOU_μσ had a non-zero mean for SAS, likely due to the small number of SAS participants in AOU.REF. The KS test was not rejected for AFR, AMR, EAS, EUR and ADM. Notably, both AD_μ and AD.AOU_μσ resulted in a mean of zero and s.d. of one for the highly admixed ADM group. Given that we are unable to perform statistical tests in the SAS cluster due to low sample size, we chose to continue with these calibrations and make comparisons between RAW, PC_μ, AD_μ, PC.AOU_μσ, and AD.AOU_μσ.

**Table 4:**
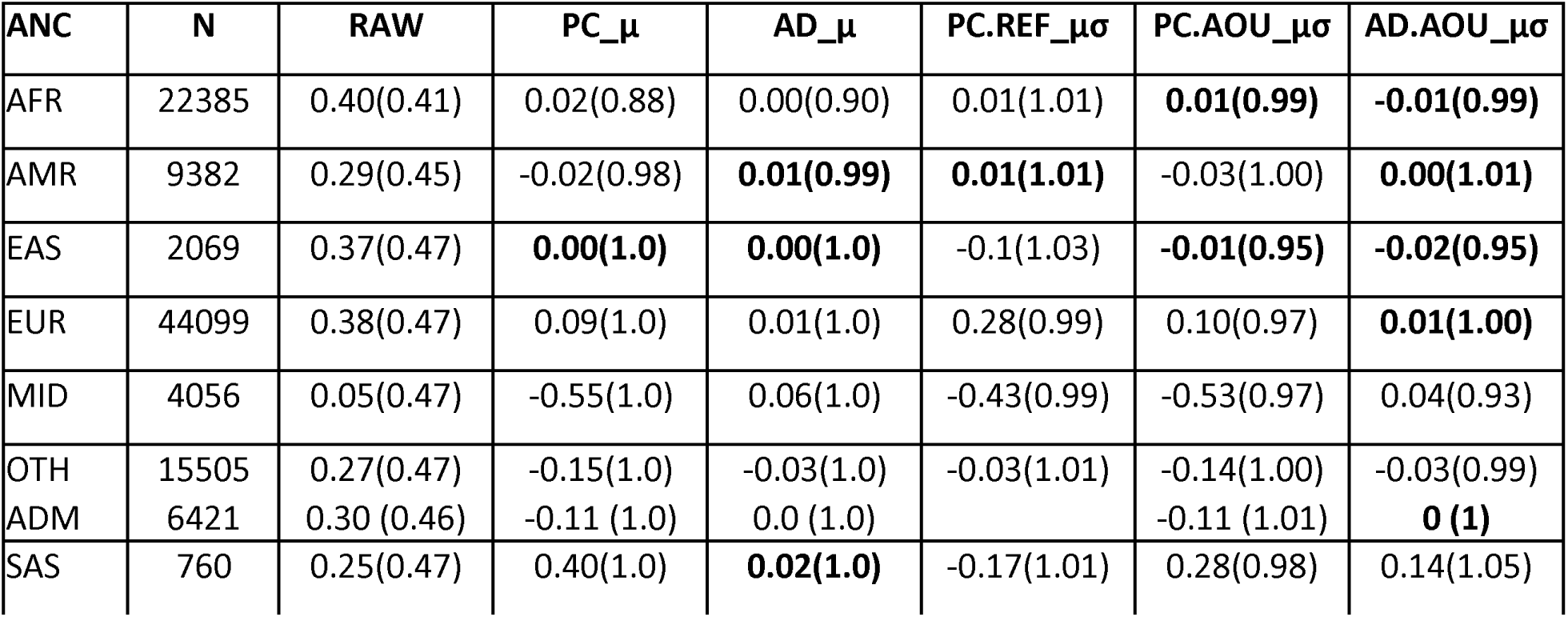
Mean (s.d.) of PRS by ancestry. Results in bold indicate for which ancestries the Kolmogorov-Smirnov test was not rejected. AFR=African, AMR=Admixed American, EAS=East Asian, EUR=European, MID=Middle Eastern, OTH=Other, ADM = Highly Admixed subset of OTH, SAS=South Asian. RAW=raw PRS; PC_μ=raw PRS adjusted for first 5 PCs of ancestry; AD_μ=raw PRS adjusted for admixture; PC.REF_μσ=raw PRS adjusted using a training model developed using 1KG and HGDP reference data. PC.AOU_μσ=raw PRS adjusted using a training model developed using a random subset of unscreened controls (AOU.REF). AD.AOU_μσ=raw PRS adjusted using a training model developed using AOU.REF and admixture estimates rather than PCs.

| ANC | N | RAW | PC_μ | AD_μ | PC.REF_μσ | PC.AOU_μσ | AD.AOU_μσ |
| --- | --- | --- | --- | --- | --- | --- | --- |
| AFR | 22385 | 0.40(0.41) | 0.02(0.88) | 0.00(0.90) | 0.01(1.01) | **0.01(0.99)** | **-0.01(0.99)** |
| AMR | 9382 | 0.29(0.45) | -0.02(0.98) | **0.01(0.99)** | **0.01(1.01)** | -0.03(1.00) | **0.00(1.01)** |
| EAS | 2069 | 0.37(0.47) | **0.00(1.0)** | **0.00(1.0)** | -0.1(1.03) | **-0.01(0.95)** | **-0.02(0.95)** |
| EUR | 44099 | 0.38(0.47) | 0.09(1.0) | 0.01(1.0) | 0.28(0.99) | 0.10(0.97) | **0.01(1.00)** |
| MID | 4056 | 0.05(0.47) | -0.55(1.0) | 0.06(1.0) | -0.43(0.99) | -0.53(0.97) | 0.04(0.93) |
| OTH | 15505 | 0.27(0.47) | -0.15(1.0) | -0.03(1.0) | -0.03(1.01) | -0.14(1.00) | -0.03(0.99) |
| ADM | 6421 | 0.30 (0.46) | -0.11 (1.0) | 0.0 (1.0) |  | -0.11 (1.01) | **0 (1)** |
| SAS | 760 | 0.25(0.47) | 0.40(1.0) | **0.02(1.0)** | -0.17(1.01) | 0.28(0.98) | 0.14(1.05) |

**Figure 1:**
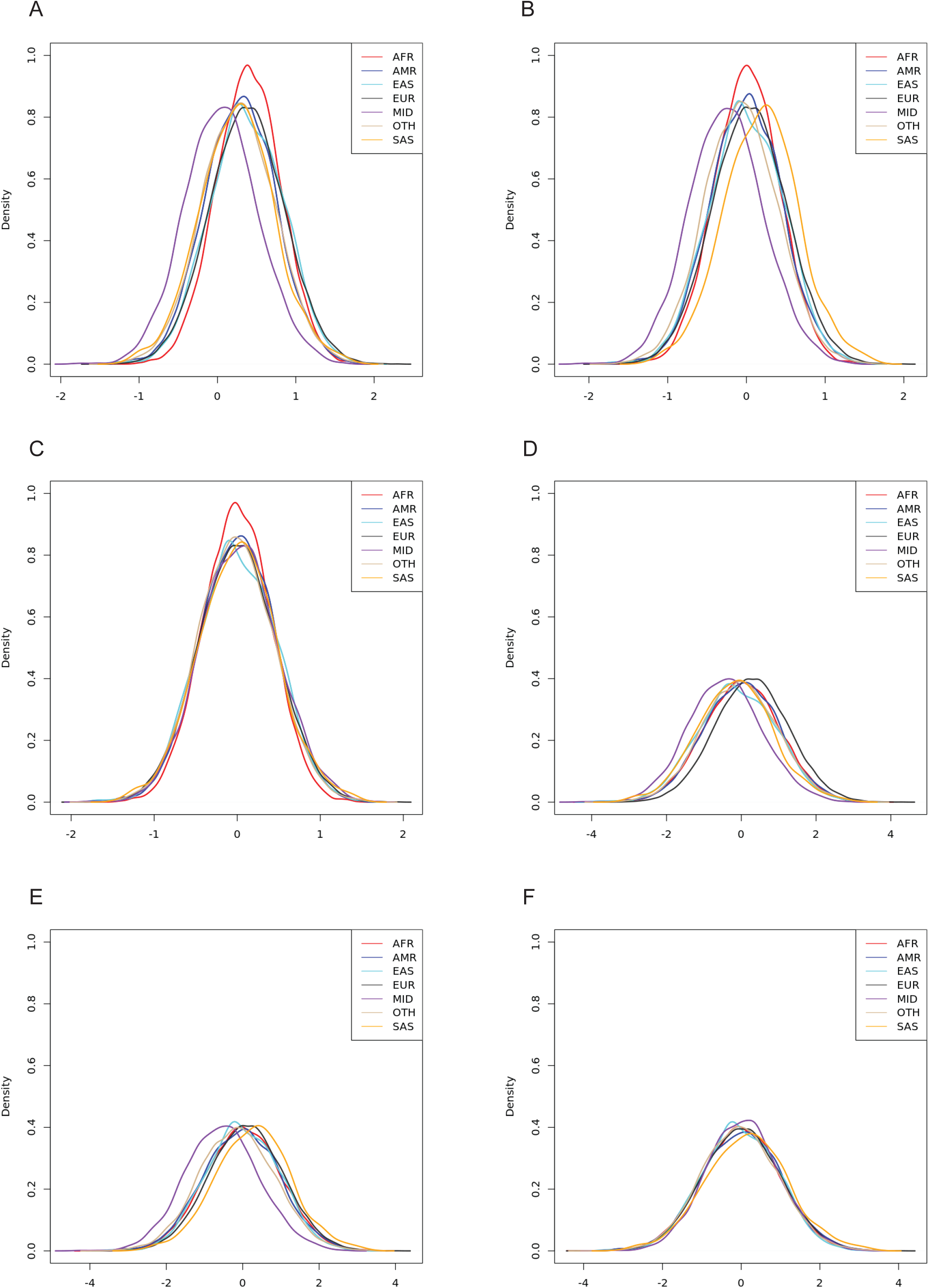
Each PRS by ancestry. A) RAW=original PRS. B) PC_μ=PRS adjusted by the first 5 PCs of ancestry in AOU. C) AD_μ=PRS adjusted by admixture estimates in AOU. D) PC.REF_μ.σ=PRS adjusted using a model trained on the first 5 PCs of ancestry in the 1KG and HGDP reference dataset. E) PC.AOU_μσ=PRS adjusted using a model trained on the first 5 PCs of ancestry in the subset of unassigned AOU participants (AOU.REF). F) AD.AOU_μσ=PRS adjusted using a model trained on admixture in AOU.REF. AFR=African, AMR=Admixed American, EAS=East Asian, EUR=European, MID=Middle Eastern, OTH=Other, SAS=South Asian.

### PRS OR

Results for the OR per 1 s.d. change in the PRS were consistent for RAW and across the calibration methods, within genetic ancestries (Table 5). The OR estimate was highest for AMR (range 2.1-2.2), was statistically significant, and had wide confidence intervals. For EUR, the estimated OR ranged between 1.6 and 1.7, was statistically significant, and had narrow confidence intervals reflecting the large sample size. For OTH, the estimated OR ranged between 1.5 and 1.6 and was statistically significant. The estimated OR for AFR ranged between 1.4 and 1.5, and was not significant after adjusting for age and sex. As this lack of significance could be due to a sex imbalance among the cases and controls, we performed sex stratified analysis in AFR. Sex stratified analysis in AFR resulted in similar estimated ORs and they were not statistically significant (data not shown). The estimated OR for MID ranged between 1.05 and 1.12, and was not statistically significant in any model. Although the ORs indicate that an increase in PRS is associated with increased risk of CRC, statistical significance for the uppermost quintile compared to the middle is observed only in EUR, due to sample size (Figures 2-6). For AMR, there were no cases in the lowest quintile.

**Table 5:** Estimated OR (95% C.I.) per s.d. change in PRS, unadjusted (first row) and adjusted for age and sex (second row), by ancestry. AFR=African, AMR=Admixed American, EUR=European, MID=Middle Eastern, OTH=Other. RAW=raw PRS; PC_μ=raw PRS adjusted for first 5 PCs of ancestry; AD_μ=raw PRS adjusted for admixture; PC.AOU_μσ=raw PRS adjusted using a training model developed using a random subset of unscreened controls (AOU.REF). AD.AOU_μσ=raw PRS adjusted using a training model developed using AOU.REF and admixture estimates rather than PCs.

| Ancestry | RAW | PC_μ | PC.AOU_μσ | AD_μ | AD.AOU_μσ |
| --- | --- | --- | --- | --- | --- |
| AFR | 1.5(1.02,2.08) | 1.5(1.02,2.07) | 1.4(1.01,1.93) | 1.4(1.01,2.04) | 1.4(1.01,1.91) |
|  | 1.4(0.99,2.02) | 1.4(0.99,2.02) | 1.4(0.99,1.88) | 1.4(0.98,1.99) | 1.4(0.98,1.87) |
| AMR | 2.2(1.27,3.85) | 2.2(1.26,3.80) | 2.2(1.26,3.72) | 2.2(1.29,3.84) | 2.2(1.28,3.72) |
|  | 2.2(1.24,3.75) | 2.1(1.23,3.70) | 2.1(1.23,3.61) | 2.2(1.26,3.74) | 2.1(1.25,3.63) |
| EUR | 1.6(1.43,1.89) | 1.6(1.43,1.88) | 1.7(1.46,1.95) | 1.6(1.43,1.87) | 1.7(1.45,1.92) |
|  | 1.7(1.44,1.90) | 1.7(1.44,1.90) | 1.7(1.47,1.96) | 1.6(1.44,1.88) | 1.7(1.46,1.93) |
| MID | 1.1(0.71,1.63) | 1.1(0.72,1.63) | 1.1(0.7,1.67) | 1.1(0.74,1.67) | 1.1(0.71,1.77) |
|  | 1.1(0.7,1.60) | 1.1(0.7,1.59) | 1.1(0.68,1.63) | 1.1(0.72,1.63) | 1.1(0.69,1.73) |
| OTH | 1.5(1.17,1.98) | 1.5(1.16,1.97) | 1.5(1.17,1.99) | 1.5(1.16,1.97) | 1.5(1.17,2) |
|  | 1.5(1.18,2.01) | 1.5(1.17,2.00) | 1.6(1.18,2.02) | 1.5(1.18,2) | 1.5(1.18,2.04) |

**Figure 2:**
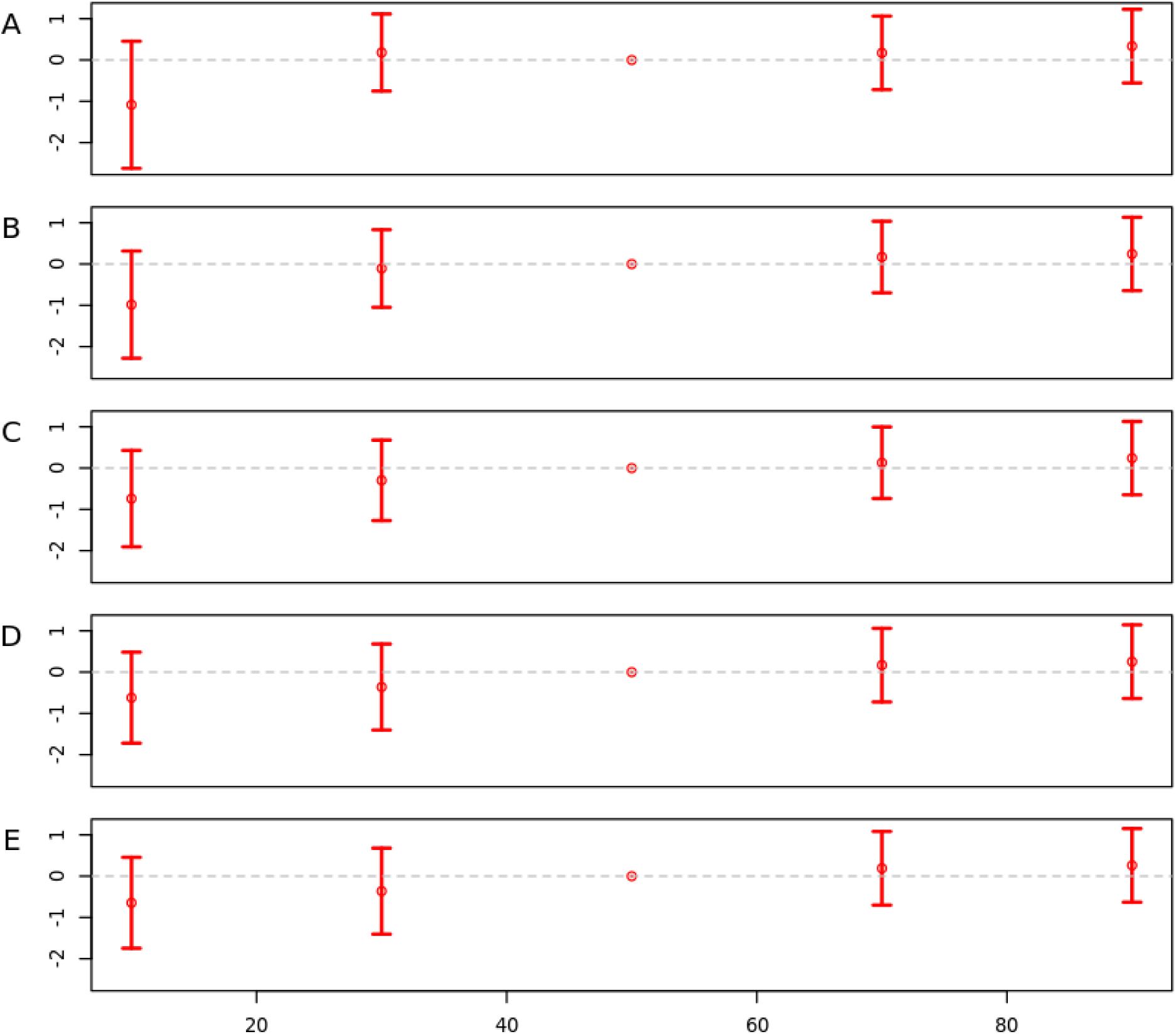
log(OR) by quintiles and calibration for African ancestry. A) RAW=raw PRS; B) PC_μ=raw PRS adjusted for first 5 PCs of ancestry; C) AD_μ=raw PRS adjusted for admixture; D) PC.AOU_μσ=raw PRS adjusted using a training model developed using a random subset of unscreened controls (AOU.REF) E) AD.AOU_μσ=raw PRS adjusted using a training model developed using AOU.REF and admixture estimates rather than PCs.

**Figure 3:**
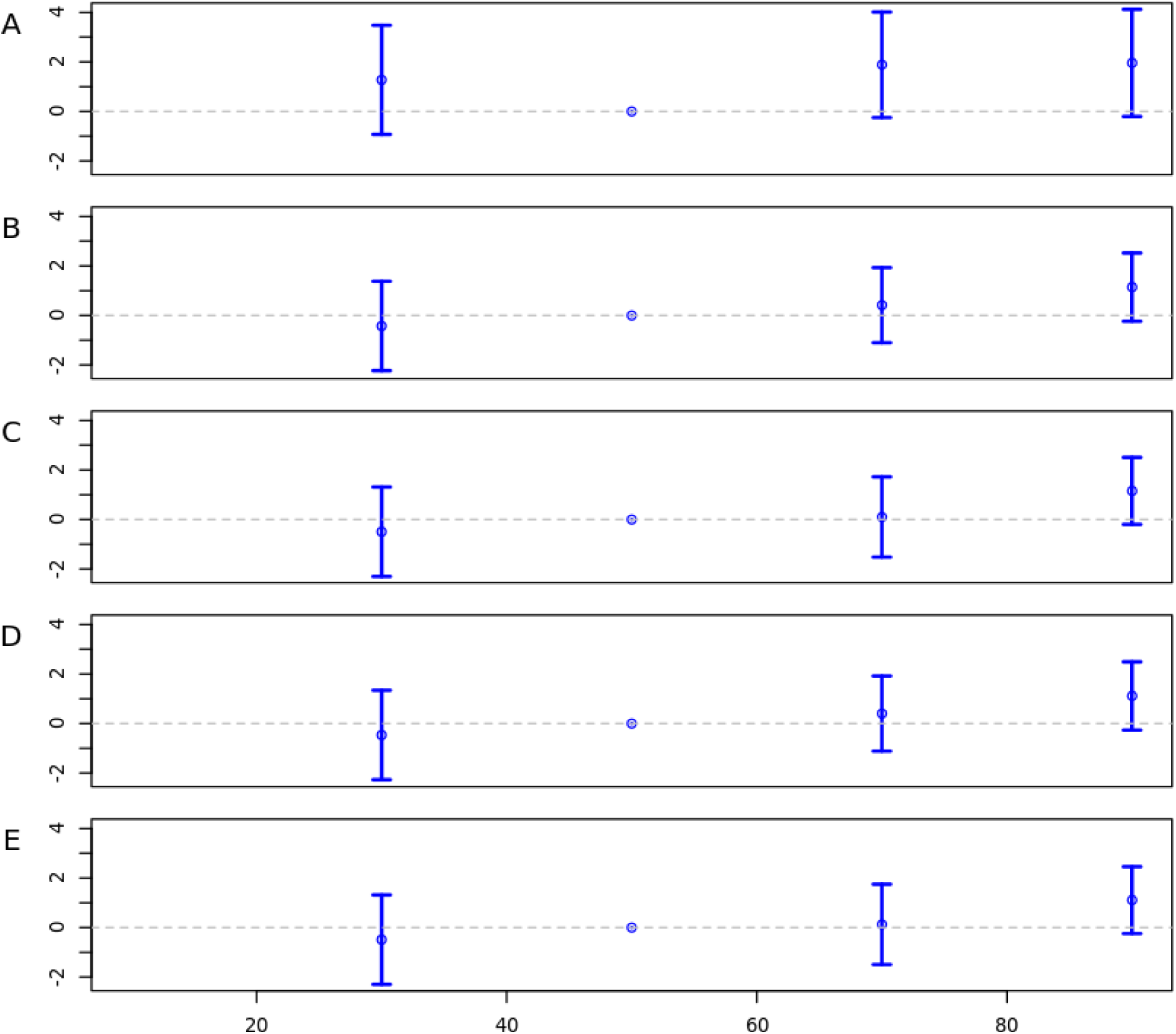
log(OR) by quintiles and calibration for Admixed American ancestry. A) RAW=raw PRS; B) PC_μ=raw PRS adjusted for first 5 PCs of ancestry; C) AD_μ=raw PRS adjusted for admixture; D) PC.AOU_μσ=raw PRS adjusted using a training model developed a using random subset of unscreened controls (AOU.REF). E) AD.AOU_μσ=raw PRS adjusted using a training model developed using AOU.REF and admixture estimates rather than PCs. OR is not calculated in the lowest quintile as there were no cases with a PRS in this quintile.

**Figure 4:**
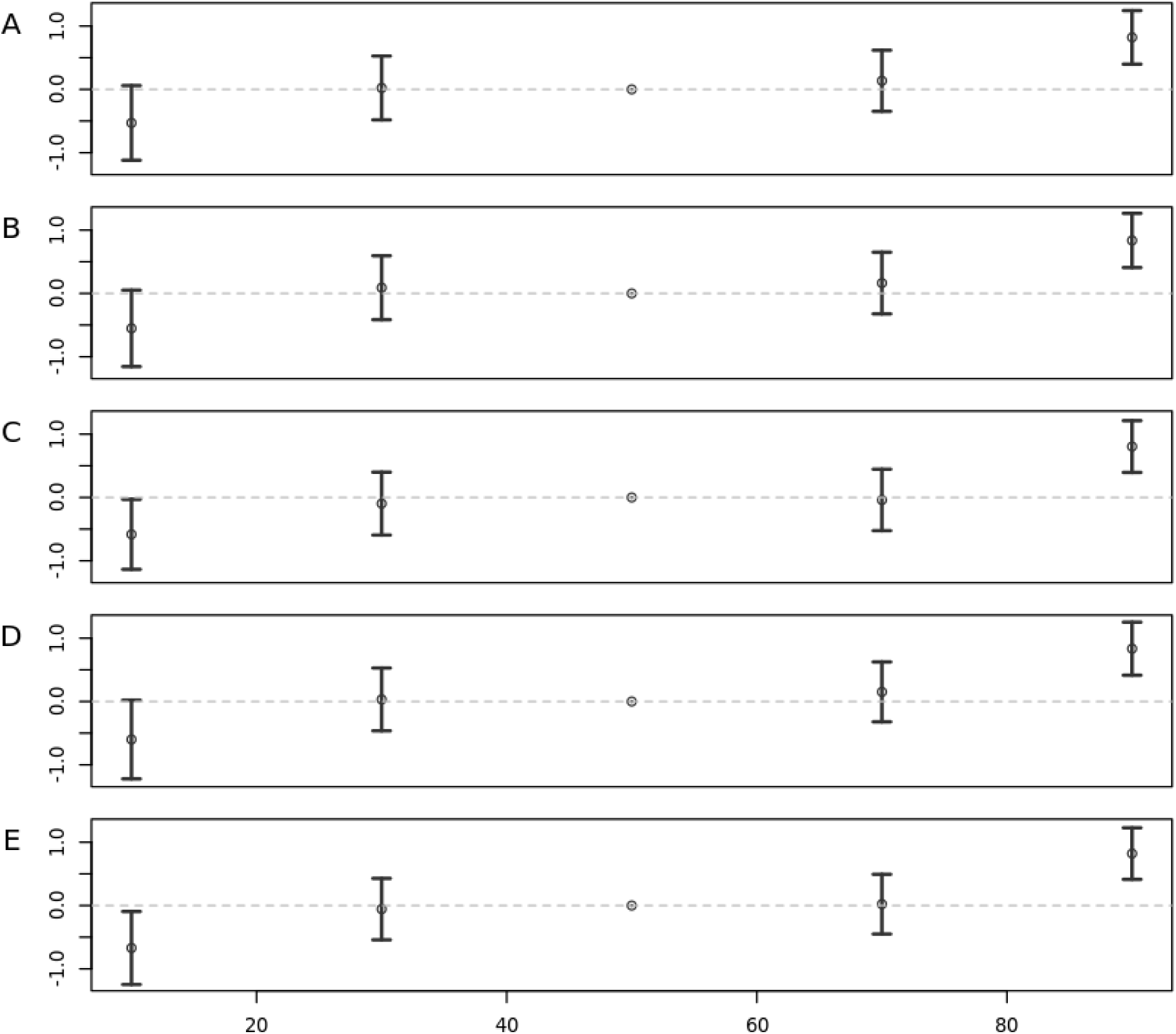
log(OR) by quintiles and calibration for European ancestry. RAW=raw PRS; PC_μ=raw PRS adjusted for first 5 PCs of ancestry; AD_μ=raw PRS adjusted for admixture; PC.AOU_μσ=raw PRS adjusted using a training model developed using a random subset of unscreened controls (AOU.REF). AD.AOU_μσ=raw PRS adjusted using a training model developed using AOU.REF and admixture estimates rather than PCs.

**Figure 5:**
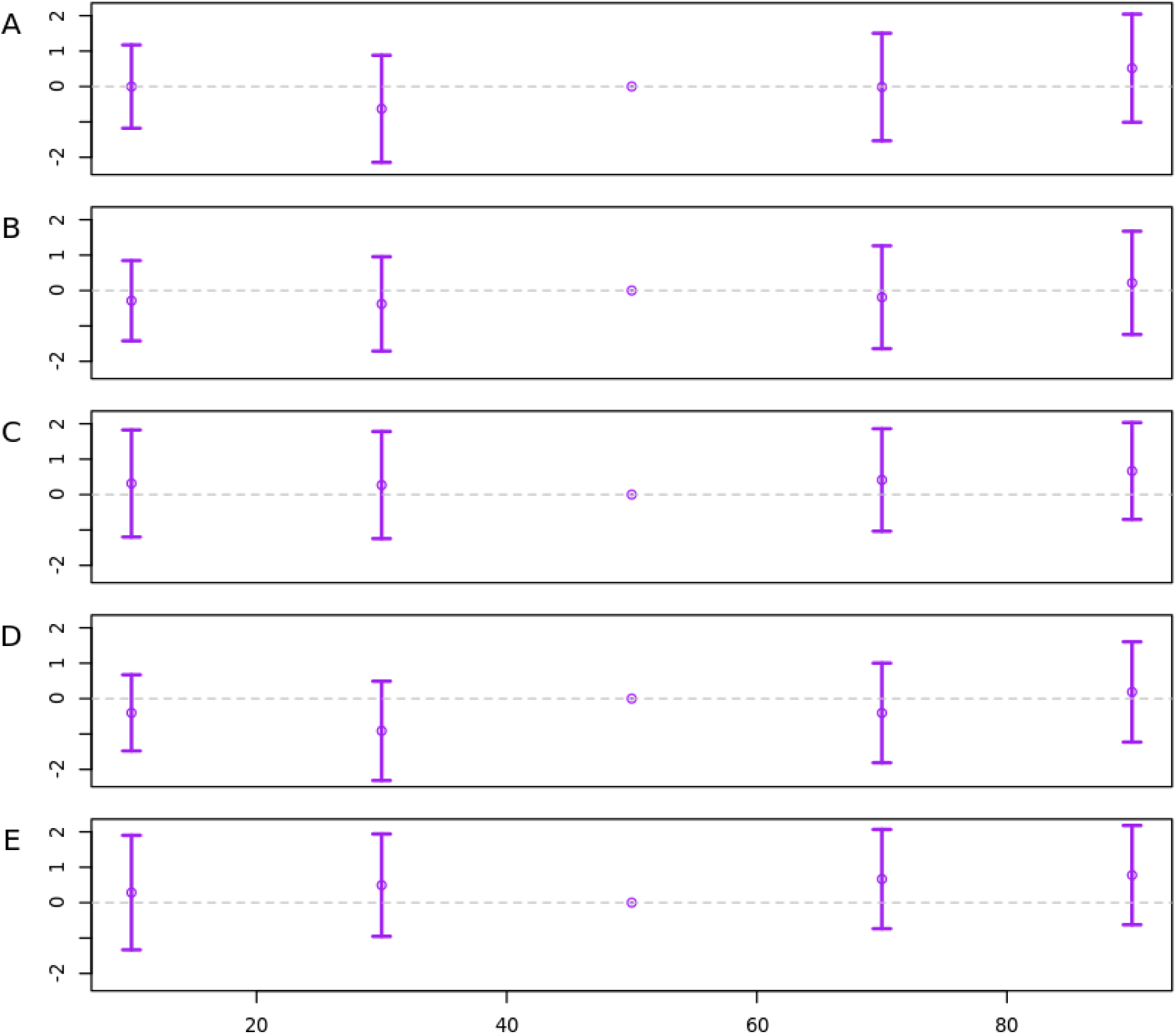
log(OR) by quintiles and calibration for Middle Eastern ancestry. A) RAW=raw PRS; B) PC_μ=raw PRS adjusted for first 5 PCs of ancestry; C) AD_μ=raw PRS adjusted for admixture; D) PC.AOU_μσ=raw PRS adjusted using a training model developed using a random subset of unscreened controls (AOU.REF). E) AD.AOU_μσ=raw PRS adjusted using a training model developed using AOU.REF and admixture estimates rather than PCs.

**Figure 6:**
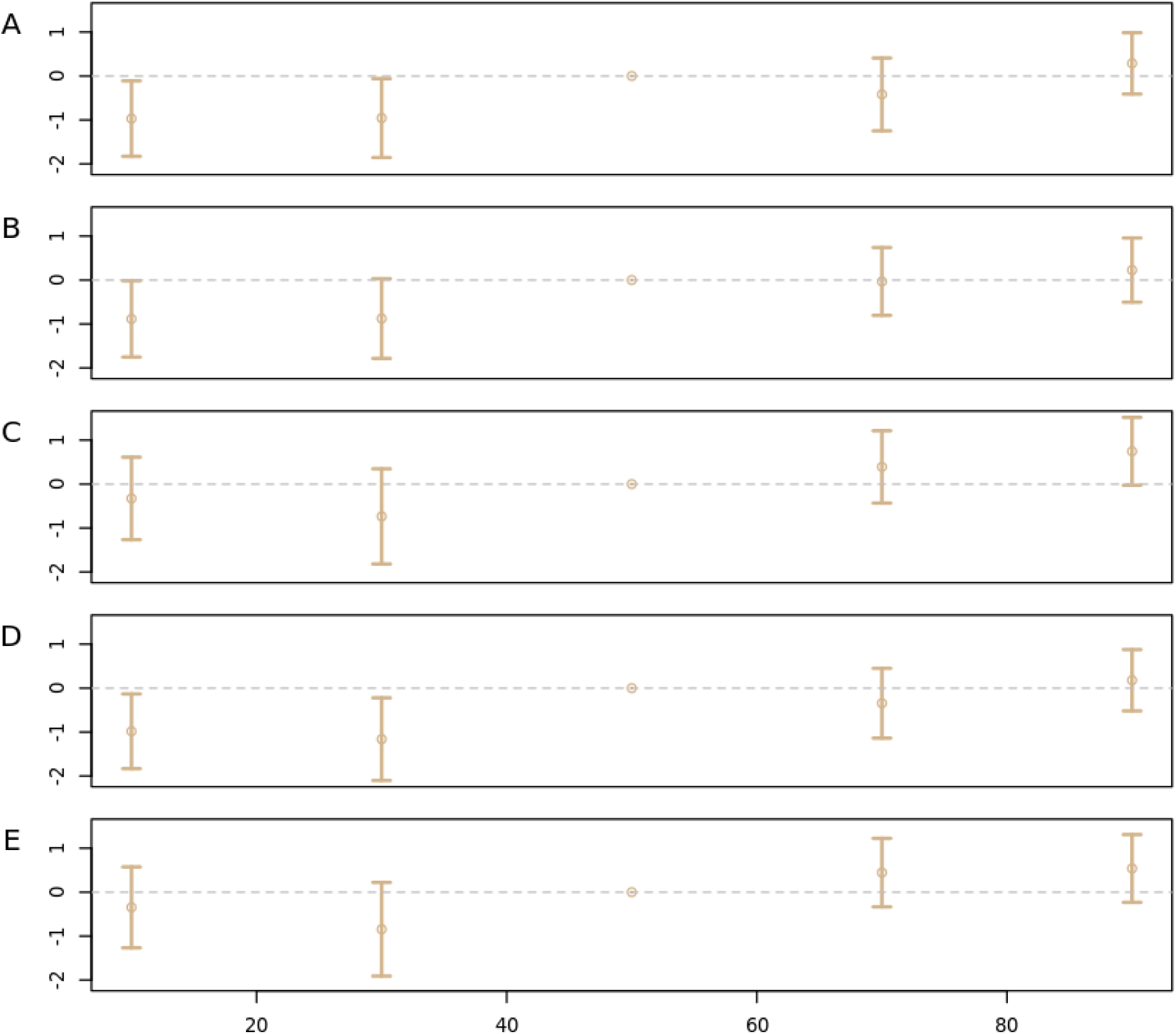
log(OR) by quintiles and calibration for Other ancestry. A) RAW=raw PRS; B) PC_μ=raw PRS adjusted for first 5 PCs of ancestry; C) AD_μ=raw PRS adjusted for admixture; D) PC.AOU_μσ=raw PRS adjusted using a training model developed using random subset of unscreened controls (AOU.REF.) E) AD.AOU_μσ=raw PRS adjusted using a training model developed using AOU.REF and admixture estimates rather than PCs.

### PRS AUC

The estimated AUC and its 95% C.I. are given in Table 6. Overall, these results are similar to those for the OR, as expected^33^. The estimated AUC is greater than 0.5 for all ancestries and all adjustments. However, the AUC is not statistically significantly different from 0.5 for both AFR for some calibrations and when adjusting for age and sex, as well as for MID in all situations. The AUC is highest for AMR (0.68-0.72), with wide C.I.s. The AUC is similar for EUR and OTH (∼0.64). EUR has the narrowest C.I. reflecting the larger sample size. Adjusting the AUC for age and sex does not change the results for AMR, EUR or OTH.

**Table 6:**
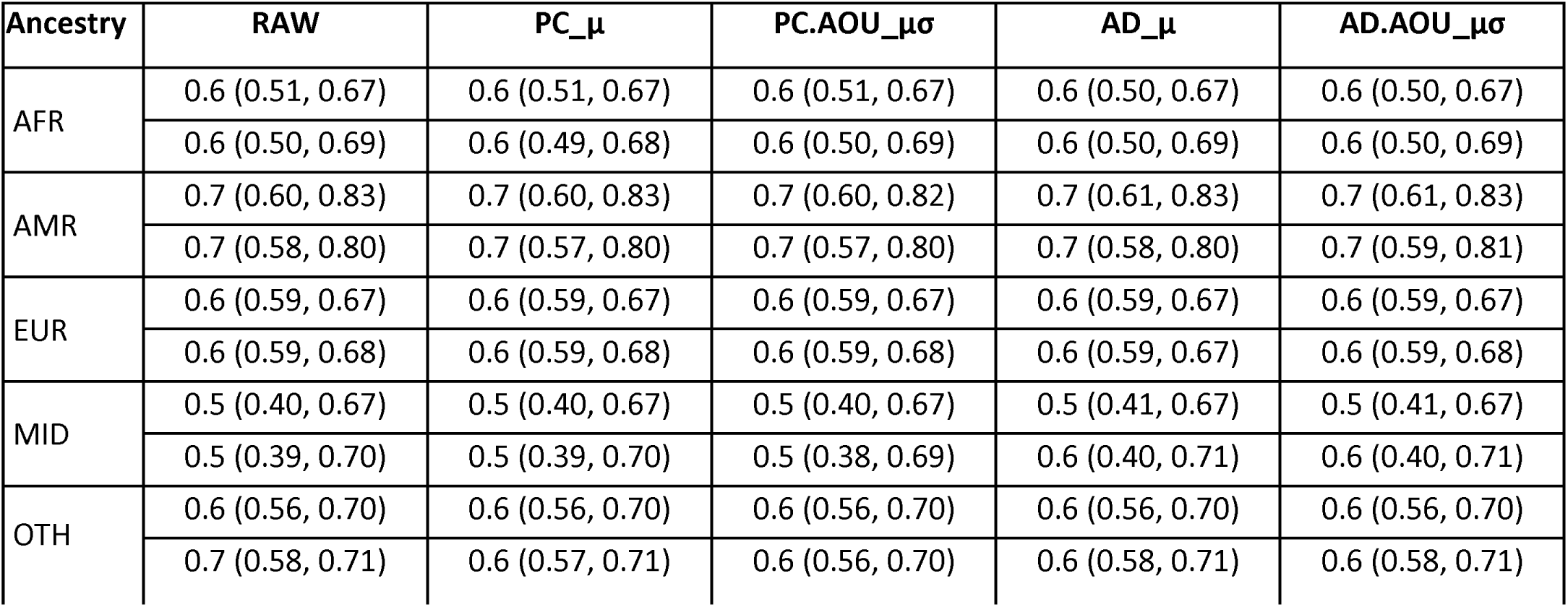
Estimated AUC and 95% C.I. for each PRS, by ancestry, comparing cleaned cases to screened controls. For each PRS, the first row contains results for the unadjusted AUC and the second row contains results for the AUC adjusted for age and sex. Although the theoretical lower bound for the C.I. is 0.5, we include the actual lower value from the bootstrap estimation. AFR=African, AMR=Admixed American, EUR=European, MID=Middle Eastern, OTH=Other. RAW=raw PRS; PC_μ=raw PRS adjusted for first 5 PCs of ancestry; AD_μ=raw PRS adjusted for admixture; PC.AOU_μσ=raw PRS adjusted using a training model developed using a random subset of unscreened controls (AOU.REF). AD.AOU_μσ=raw PRS adjusted using a training model developed using AOU.REF and admixture estimates rather than PCs.

| Ancestry | RAW | PC_μ | PC.AOU_μσ | AD_μ | AD.AOU_μσ |
| --- | --- | --- | --- | --- | --- |
| AFR | 0.6 (0.51, 0.67) | 0.6 (0.51, 0.67) | 0.6 (0.51, 0.67) | 0.6 (0.50, 0.67) | 0.6 (0.50, 0.67) |
|  | 0.6 (0.50, 0.69) | 0.6 (0.49, 0.68) | 0.6 (0.50, 0.69) | 0.6 (0.50, 0.69) | 0.6 (0.50, 0.69) |
| AMR | 0.7 (0.60, 0.83) | 0.7 (0.60, 0.83) | 0.7 (0.60, 0.82) | 0.7 (0.61, 0.83) | 0.7 (0.61, 0.83) |
|  | 0.7 (0.58, 0.80) | 0.7 (0.57, 0.80) | 0.7 (0.57, 0.80) | 0.7 (0.58, 0.80) | 0.7 (0.59, 0.81) |
| EUR | 0.6 (0.59, 0.67) | 0.6 (0.59, 0.67) | 0.6 (0.59, 0.67) | 0.6 (0.59, 0.67) | 0.6 (0.59, 0.67) |
|  | 0.6 (0.59, 0.68) | 0.6 (0.59, 0.68) | 0.6 (0.59, 0.68) | 0.6 (0.59, 0.67) | 0.6 (0.59, 0.68) |
| MID | 0.5 (0.40, 0.67) | 0.5 (0.40, 0.67) | 0.5 (0.40, 0.67) | 0.5 (0.41, 0.67) | 0.5 (0.41, 0.67) |
|  | 0.5 (0.39, 0.70) | 0.5 (0.39, 0.70) | 0.5 (0.38, 0.69) | 0.6 (0.40, 0.71) | 0.6 (0.40, 0.71) |
| OTH | 0.6 (0.56, 0.70) | 0.6 (0.56, 0.70) | 0.6 (0.56, 0.70) | 0.6 (0.56, 0.70) | 0.6 (0.56, 0.70) |
|  | 0.7 (0.58, 0.71) | 0.6 (0.57, 0.71) | 0.6 (0.56, 0.70) | 0.6 (0.58, 0.71) | 0.6 (0.58, 0.71) |

### Observed Upper Percentiles

We compared the observed upper 5, 7.5 and 10^th^ percentiles to their expected values for each PRS. (Figure 7). The 95% C.I.s for RAW are below expected risks for AFR, AMR, MID, and OTH, and above expected risks for EUR, indicating that RAW overestimates the number of EUR who are at high risk and underestimates the numbers from the other ancestries. The 95% C.I.s for PC_μ are below the estimated risks for AFR, AMR, MID, and OTH and above the estimated risks for EUR, similar to RAW. The 95% C.I.s for PC.AOU_μσ covers the expected risks for AFR, covers the expected risk for AMR at the 5^th^ percentile only, are below the expected risks for AMR (7.5, 10^th^ percentiles), MID, and OTH, and above the estimated risks for EUR, indicating that PC.AOU_μσ correctly estimates the number of high risk patients in AFR and AMR (5^th^ percentile), but over/under-estimates them in the other ancestry clusters. The 95% C.I.s for AD_μ cover the expected risks for AMR and OTH, are below the expected risks for AFR and above the expected risks for EUR, indicating that AD_μ correctly estimates the number of high risk patients for AMR and OTH, but not the other ancestires. The 95% C.I.s for AD.AOU_μσ contains the expected risks for AMR, EUR, and MID, and are just below the expected risks for AFR and OTH, indicating that AD.AOU_μσ correctly estimates the number of high risk patients in AMR, EUR, and MID, and slightly understimates them for AFR and OTH. In the highly admixed group, ADM, the 95% C.I.s include the expected risks for only AD_μ and AD.AOU_μσ, indicating that only these two calibration methods correctly estimate the number of high risk patients among the highly admixed participants. We do not comment on the risk assessment for EAS and SAS due to the large C.I. in these clusters, reflecting their smaller sample sizes.

**Figure 7:**
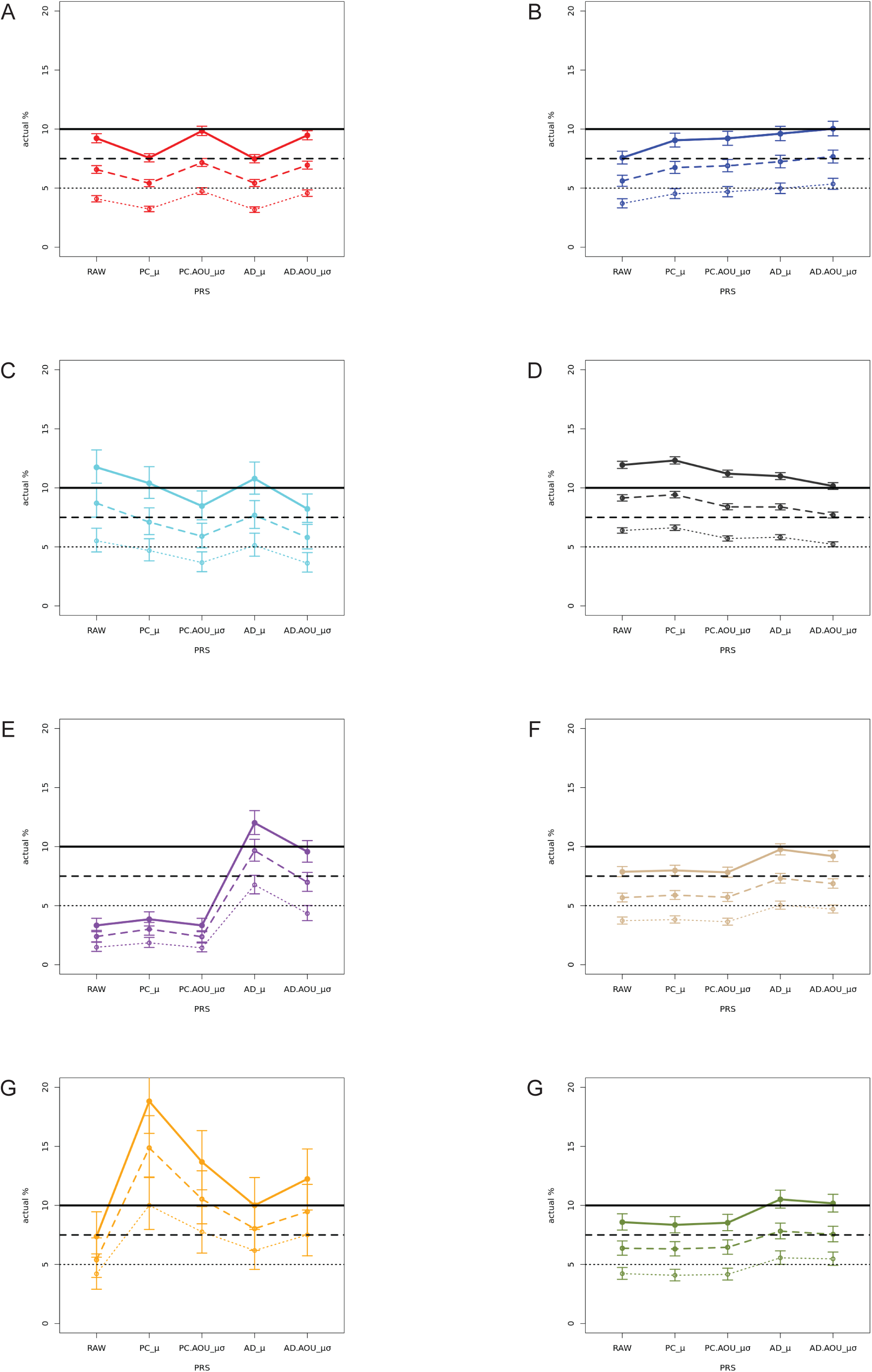
Observed versus expected upper percentiles, by genetic ancestry and PRS. A) African B) Admixed American C) East Asian D) European E)Middle Eastern F) Other G) South Asian, and H) Highly Admixed (subset of Other). For each panel, solid colored line is observed upper 10th percentile. Dashed colored line is observed upper 7.5 percentile. Dotted colored line is observed upper 5th percentile. 95% C.I. are indicated with error bars. Horizontal black solid, dashed and dotted lines indicate the expected percentiles, respectively. RAW=raw PRS; PC_μ=raw PRS adjusted for first 5 PCs of ancestry; AD_μ=raw PRS adjusted for admixture; PC.AOU_μσ=raw PRS adjusted using a training model developed using a random subset of unscreened controls (AOU.REF). AD.AOU_μσ=raw PRS adjusted using a training model developed using AOU.REF and admixture estimates rather than PCs.

## Discussion

Regardless of the ancestry calibration, the PRS was statistically significantly associated with CRC status for EUR, AMR, and OTH, was borderline significant for AFR, and was not significant for MID. Additionally, the strength of the significant effects differed across ancestries. The estimated OR and AUC were highest for AMR, possibly due to the lack of cases in the lowest quintile of the PRS. In addition, the confidence interval for AMR was large, as the total sample size was small. The OR estimate was higher in EUR than in OTH, but they had similar AUC. The sex imbalance between cases and controls in AFR may contribute to the borderline significance observed in that cluster, but sex stratified analysis did not have enough power to confirm this, due to low sample size.

Power was sufficient in only the large EUR cluster to show that participants with PRS in the upper quintile were at higher risk of CRC than those at typical risk (the middle quintile). Observed differences in OR and their large C.I. comparing quintiles in other ancestries is due to the low number of cases within each quintile, and the overall smaller sample sizes within each quintile. Specifically, we did not observe an OR for the lowest quintile in AMR, as there were no cases with a PRS in that quintile.

Although the predictiveness of the association between the PRS and risk is not affected by genetic ancestry calibration, within ancestral groups, the resulting distributions are affected, which can influence downstream clinical decisions or overall risk estimation when incorporating environmental and lifestyle factors. Adjusting for the first 5 PCs of ancestry resulted in non-zero means for some ancestries, whereas adjusting for admixture estimates resulted in means close to zero for all ancestries (except SAS)The AD.AOU_μσ calibration method resulted in a standard Normal distribution for the highly admixed cohort, as well as AFR, AMR, EAS and EUR, and was close to standard Normal for all other ancestries (except SAS). Finally, the upper tail frequencies were closely or accurately estimated by AD.AOU_μσ only.

There are several limitations to this study. The sample sizes for EAS and SAS are too small to make any inferences about those genetic clusters. Furthermore, the availability of EHR data, and therefore the number of cases and controls, may be associated with sex and self-identified race or ancestry ^25^. This may affect the power to detect associations in the AFR genetic cluster as well as when comparing the upper quintiles to the middle quintile for AFR, AMR, and OTH. Additionally, the estimated effect sizes observed in this study may be an underestimate as cases in AOU data may represent a healthier cohort than typical CRC cases due to survival bias. This is evidenced by the fact that the diagnosis of CRC occurred before entry into the study by a mean of 5 years with a maximum of 27 years. This phenomenon of biobank participants being healthier than the population they represent has been observed previously ^34,35^.

Another limitation is with the PRS, itself. Although the PRS is multi-ancestry, it was developed using participants from only two global populations: EUR and EAS. It has been shown that the applicability of PRS declines with further genetic ancestry distance from the population used to develop a PRS ^36^, which we observe here. Continued development of the PRS, incorporating samples from other ancestral populations should resolve the differences in predicted accuracy by genetic ancestry. As we used whole genome sequence data, observed differences in OR across ancestries cannot be attributed to uncertainty in imputation of genotypes, but rather are due to differences in MAF, LD with causal variation, and possible effect heterogeneity across ancestries. Interestingly, we observe the highest OR and AUC for AMR, which appears to cluster between EUR and EAS in PC1xPC2 space. However, this phenomenon has yet to be analyzed for PRS that are developed in more than one population, as in this study.

Some methodological choices also limit the study. The global populations used to calculate the PCs of ancestry may be too general and not comprehensive. For example, we defined AFR ancestry as those that cluster with reference samples from East and West Africa, the Southwest U.S. and the Caribbean, which encompasses a large swath of diversity. Similarly, the reference samples that made up the MID cluster were limited to a small region that included samples from Druze, Moabite, Palestinian and Bedouin populations. We tried to accommodate these differences by allowing for more admixture among the AFR, AMR and MID clusters. Furthermore, the global ancestry references did not contain Pacific Islander or Native American samples, so we are unable to analyze the applicability of the PRS or the *post-hoc* calibration in these populations.

Another methodological choice was the size of the reference patient sample. We chose to limit the sample size to be the same as that in the global reference sample. However, a larger sample size for the patient reference may have resulted in improved training models for PC.AOU_μσ and AD.AOU_μσ for both the mean and variance corrections, as well as the resulting distributions being closer to standard Normal. Additionally, the sampling method could be further improved to ensure that the PC space is adequately sampled and the resulting patient reference sample represents the patient population of interest. Finally, the availability of millions of SNP genotypes for both PC projection and PRS calculation might be a barrier. However, the increased availability of WGS and genotype imputation based on widely available SNP chips should relieve this issue.

A comprehensive score which combines a calibrated multi-ancestry PRS with social determinants of health, and other known risk factors, is the ultimate goal for use in the clinic. We acknowledge that further development of the multi-ancestry PRS is necessary for equitable risk prediction across genetic ancestry. However, an improved raw PRS will still be associated with genetic ancestry due to LD and MAF differences across ancestries. We believe that a *post-hoc* genetic ancestry calibration based on the AD.AOU_μσ method will be most effective as it generates distributions closest to standard Normal for all genetic backgrounds, can be applied to any patient, and does not require the clinician to determine the patient’s ancestry.

## Acknowledgments

The authors would like to thank Alyna T. Khan for her valuable comments and suggestions. This work was funded by the Office of the Director at the National Institute of Health, under award notice 1OT2OD002748-01 and by the NHGRI through the grant U01HG008657.

## Data and code availability

Data from the NIH All of Us study are available via institutional data access for researchers who meet the criteria for access to confidential data. To register as a researcher with All of Us, researchers may use the following URL and complete the laid out steps: https://www.researchallofus.org/register/. Researchers can contact All of Us Researcher Workbench Support at. Procedures were followed in accordance with the ethical standards of the Institutional Review Board (IRB) of the All of Us Research Program. The All of Us IRB follows the regulations and guidance of the NIH Office for Human Research Protections for all studies, ensuring that the rights and welfare of research participants are overseen and protected uniformly. Proper informed consent was obtained. Before signing forms saying that they consent to participate, participants view a series of screens with text and short videos that explain the program’s goals, how it works, and what participation entails. Code used in this study is available at the Researcher Workbench “Compare ancestry calibration methods for a CRC PRS” located at https://workbench.researchallofus.org/workspaces/aou-rw-ed3b00c1/compareancestrycalibrationmethodsforacrcprs/analysis.

## Web Resources

Online Mendelian Inheritance in Man: http://www.omim.org

Colorectal Cancer case/control algorithm: phekb.org/phenotype/colorectal-cancer-crc CRC PRS: https://www.pgscatalog.org/score/PGS003852/

1000 Genomes Data: https://www.internationalgenome.org/home

https://www.internationalgenome.org/data-portal/search?q=1000%2Bgenomes

Human Genome Diversity Project: https://www.internationalgenome.org/data-portal/data-collection/hgdp

