## Supplemental Methods for "Comparing ancestry calibration approaches for a trans-ancestry colorectal cancer polygenic risk score"

#### Principal Components and Admixture

We generated Principal Components of Ancestry using a set of ancestry informative markers from the Illumina Infinium Global Diversity Array-8[1]. We used HAIL [2] on the Spark cluster to extract genotypes from whole genome sequence (WGS) data at these markers in both the reference 1000 Genomes (1KG) and Human Genome Diversity Project (HGDP) (N=4,151) and the All of Us (AOU) participants (N=98,622). We performed quality control (QC) with PLINK v1.90b6.22 [3] on the joint data as follows: First, filtered out markers with a minor allele count < 100 and genotype missingness > 0.001. Then removed SNPs with more than two alleles. We further randomly pruned the SNPs to have genotype correlation ($r^2$) < 0.2, leaving 300,387 SNPs genomewide for principal component analysis and admixture estimates.

We estimated kinship (K) among the reference genomes and randomly removed all but one individual from each related group ($K<2^{-4.5}$; first cousins or more related), leaving N=3,440 unrelated reference genomes before calculating the PCs of ancestry with the SNPRelate V1.32.1 package [4,5]. The variance explained by the first ten PCs is shown in supplemental table 1. We then projected both the remaining 711 reference samples and the participants from AOU onto the same PC space (Supplemental Figure 1).

We estimated admixture using EIGMIX and the reference global populations represented by the 1KG and HGDP unrelated set [6] (Supplemental Figure 2). The ancestry groups were defined by the continental groups: African (AFR,N=648), Admixed American/Latino (AMR,N=371), East Asian (EAS,N=703), European (EUR,N=655), Middle Eastern (MID, N=118) and South Asian (SAS, N=651), ignoring the remainder that were missing an assigned continental group (N=294). All calculations were performed in R v4.2.2 using the {bigSNPR} package v. 1.10.8.

#### PRS Calculation

We obtained the weight of each SNP calculated as described in Thompson et. al. (under review)[7], including the chromosome, position, allele 0 (a0), allele 1 (a1) and effect of the a1 allele ($\alpha$). This PRS was developed with individuals of EUR and EAS ancestry. For each individual, the PRS is calculated as the sum of the count of a1 alleles multiplied by its effect for all SNPs ($PRS = \sum_{i=1}^{K} \alpha_i G_i$), where $G_i$ is the count of a1 at SNP i and K=1,016,596. After QC (see below) in the AOU and 1KG and HGDP datasets, 1,013,380 SNPs (99.7%) of the original SNPs were used in the calculation of the PRS.

The PRS was developed on human build GRCh37. However, the whole genome sequencing data for this analysis was aligned and genotyped on human build GRCh38. We used the liftOver tool to calculate the positions on human build GRCh38 [https://genome-store.ucsc.edu/]. Of the original 1,016,596 SNPs, 376 (0.04%) did not map over from human build GRCh37 to human build GRCh38.

We used HAIL on the Spark cluster to extract genotypes from the AOU WGS and the 1KG and HGDP reference data, at the human build GRCh38 positions and output the data into PLINK format. We excluded indels as well as variants with genotype quality < 20 or genotype call rate < 10%. We further excluded participants with <90% genotype call rate. This resulted in a loss of N=3,216 SNPs in AOU and N=3,336 SNPs in the 1KG and HGDP reference data.

The PRS was calculated using the R package {bigSNPR} v. 1.10.8. Any records whose alleles did not match both a0 and a1 were assigned an effect of 0. Some SNVs were sequenced on the opposite strand or the reverse of the strand used in the original PRS derivation (e.g., alleles A and G versus T and C). We took this into account and transformed the effect size appropriately using the snp_match() command. In AOU, 2240 (0.2%) positions were sequenced in the opposite direction (reverse), 536 (0.05%) were sequenced on the other strand (flipped) and of these, 35 were sequenced on the opposite strand and in the reverse direction. In the 1KG and HGDP genome reference data, 2,239 (0.2%) positions were sequenced in the opposite direction, 536 (0.05%) were sequenced on the other strand, and of these 35 were sequenced on the opposite strand and in the reverse direction. Missing genotypes at individual SNPs were replaced with the most frequent genotype at that SNP using snp_fastImputeSimple(method="mode").

### Supplemental Tables

Table S1: Percent variance explained for the first 10 principal components of ancestry using unrelated samples from 1000 Genomes and HGDP reference data.

| Eig | 1 | 2 | 3 | 4 | 5 | 6 | 7 | 8 | 9 | 10 |
|---|---|---|---|---|---|---|---|---|---|---|
| %var | 5.37 | 2.41 | 0.82 | 0.61 | 0.27 | 0.23 | 0.17 | 0.16 | 0.14 | 0.12 |

Tables S2-S10 are available in the Supplemental spreadsheet

#### Table S2: CRC Condition Codes

Concept IDs used to determine which participants have CRC codes recorded in their electronic health record. All participants with these codes are considered to be a potential case

#### Table S3: Lynch and HNPCC Condition Codes

Concept IDs used to determine which participants have Lynch or HNPCC codes recorded in their electronic health record. These participants are excluded from the study.

#### Table S4: CRC Surgical Procedure Codes

Concept IDs used to determine which participants have had a surgery related to CRC within 12 months of diagnosis for CRC. These are used to identify true cases.

#### Table S5: Chemo and Radiation Procedure Codes

Concept IDs used to determine which participants have had chemotherapy or radiation within 12 months of CRC diagnosis. These are used to identify true cases.

#### Table S6: Chemo and Radiation Drug Codes

Concept IDs used to determine which participants have had chemotherapy or radiation drugs within 12 months of CRC diagnosis. These are used to identify true cases.

#### Table S7: non-CRC cancer Condition Codes

Concept IDs used to determine which participants have a cancer other than CRC. These participants are excluded from the analysis.

#### Table S8: Colonoscopy and Endoscopy Procedure Codes

Concept IDs used to determine which participants without CRC have had a colonoscopy or endoscopy procedure. These are used to identify screened controls.

#### Table S9: Stool Screening Measurement Codes

Concept IDs used to determine which participants without CRC have a positive stool test. These participants are excluded from analysis. In addition to these codes, pathology reports with Observational Medical Outcomes Partnership (OMOP) ID 40664885 and Stool screening positive OMOP ID 46270738 were used to exclude non-case participants from the analysis.

#### Table S10: Stool Screening Condition Codes

Concept IDs used to determine which participants without CRC have a stool screening test.

### Supplemental Figures

#### Figure S1:

The genetic ancestral groups mapped to the first two principal components of ancestry in All of Us participants. A) plot showing just the anchor ancestry clusters (AFR, EUR, EAS); B) Plot with all ancestry clusters. The EUR cluster contains the most participants, but is hidden by the other clusters.

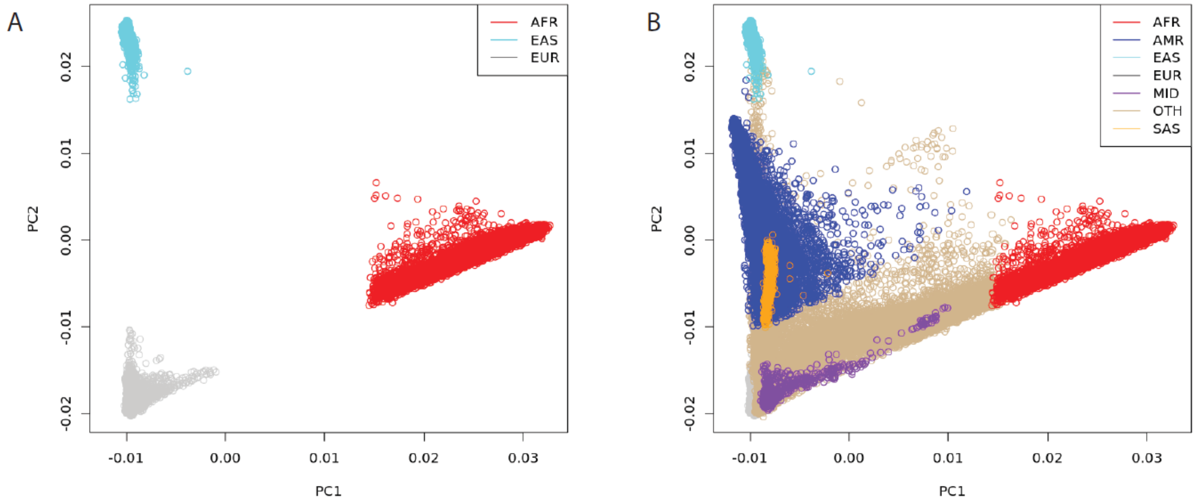

#### Figure S2:

Distribution of non-zero admixture estimates within the All of Us participants for each genetic ancestry: A) African B) Admixed American C) East Asian D) European E) Middle Eastern F) South Asian. Note that the y-axis is not the same for each figure, and represents the numbers of participants that have non-zero admixture estimates for each global ancestry used in the EIGMIX model.

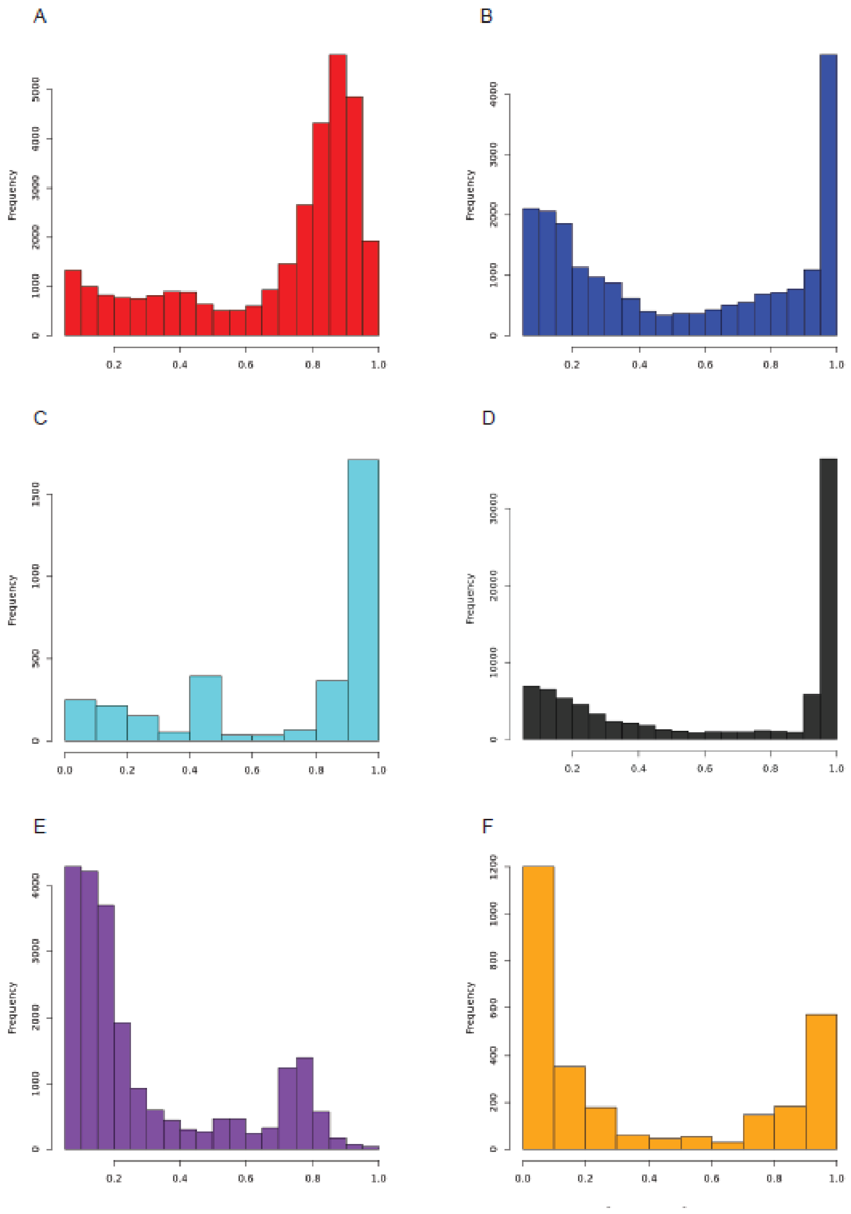
